# Predicting Prolonged Length of Stay For Hospitalized Stroke Patients Using Common Physiological Features

**DOI:** 10.1101/2024.01.21.24301586

**Authors:** Zachary Murphy, Michael Ainsworth, Kirby Gong, Elizabeth K. Zink, Joseph L. Greenstein, Raimond L. Winslow, Mona N. Bahouth

**Affiliations:** Department of Biomedical Engineering, Johns Hopkins University; Johns Hopkins University School of Medicine; Neuroscience Critical Care Unit, Johns Hopkins Medicine; Institute for Computational Medicine, Johns Hopkins University; Department of Neurology, Johns Hopkins University School of Medicine

## Abstract

**Background and Purpose:** Stroke is a leading cause of death and disability worldwide. Predicting which patients are at risk for a prolonged length of stay (LOS) could assist in coordination of care and serve as a rough measure of clinical recovery trajectory. During the acute stroke period, there is a disruption in the fidelity of the blood-brain barrier and cerebral autoregulation, and we hypothesize that trends in physiologic parameters early in a patient’s hospital course may be used to predict which patients are increased risk for a prolonged LOS. In this work we sought to create a model to predict prolonged LOS (defined as ≥ 7 days) from patient data available at admission as well as routinely collected physiologic (pulse, blood pressure, respiratory rate, temperature), and other data from the first 24 hours of admission.

**Methods:** This retrospective cohort study included stroke patients admitted to an urban comprehensive stroke center between 2016-2019. Data included common physiological parameters (pulse, temperature, blood pressure, respirations, and oxygen saturation) as well as demographic and comorbidity data. Raw time series data were transformed into statistical features for modeling. Logistic regression, random forest, and XGBoost models were trained on data collected during the first 24 hours after hospital admission to predict prolonged LOS and evaluated on a held-out test set.

**Results:** A total of 2,025 patients were included. Using an XGBoost classifier we obtained a ROC AUC of 0.85 and Precision-Recall AUC of 0.77, with the optimal operating point achieving an accuracy of 0.80, sensitivity of 0.78, specificity of 0.81.

**Conclusions:** The model suggests that prolonged LOS can be predicted with reasonable accuracy using clinical data obtained within the first 24 hours of hospitalization. This approach could provide the basis for development of a risk score and augment the care coordination process.

## INTRODUCTION

Worldwide, stroke is the second leading cause of death and the third leading cause of disability-adjusted lost years due to cognitive and physical impairments common to the disease [1]. Acute care in a stroke unit plays a major role in improving outcomes and reducing common post-stroke complications like secondary brain injury, increased intracranial pressure, seizures, cardiac complications, development of venous thromboembolism, infections, and falls [2, 3]. This standardized approach to care has led to shortened length of hospital stay by expediting diagnostic testing and prioritizing the plan of care to focus on issues that may threaten cerebral perfusion. Predicting which patients are at risk for a prolonged length of stay could 1) assist in coordination of care, and 2) serve as a rough measure of clinical recovery trajectory.

Cerebral autoregulatory dysfunction after stroke is a major concern necessitating close monitoring of blood pressure to ensure adequate cerebral blood flow and stable or improved clinical neurologic function. In the first hours and days after stroke further injury to the brain can occur due to loss of integrity of the blood brain barrier resulting in cerebral edema inflammation and disruption of cerebral blood flow associated with extension of brain tissue injury. All these processes have the potential to cause complications and worsen neurological function [2, 4]. Maintenance of conditions promoting adequate cerebral blood flow such as judicious management of hemodynamics is crucial in the survival of brain tissue and by extension the patient outcome [5]. Due to the importance of hemodynamics after a stroke, it is expected that hemodynamic parameters may be an important variable in predicting recovery trajectory and patients at risk for prolonged length of stay.

In this work we created a model to predict prolonged length of stay (defined as ≥ 7 days) from patient data available at admission as well as routinely collected physiologic data (pulse, blood pressure, respiratory rate, temperature) laboratory values, medications administered, fluid intake/output, neurologic assessments, and hospital unit assignment from the first 24 hours of admission.

## MATERIALS AND METHODS

This study was approved by the Johns Hopkins Medicine Institutional Review Board with a waiver of informed consent. The data supporting the findings of this study are available from the corresponding author upon reasonable request. All code used in this project is publicly available at https://github.com/BLINDED FOR REVIEW. This work adhered to the STROBE-RECORD reporting guidelines.

### Data source

This is a retrospective cohort study using patient data collected through the Electronic Health Record (EHR) at a single urban comprehensive stroke center between 2016 and 2019. Data was extracted and de-identified from the EHR in use at the study site, including patient demographics, comorbidities, discharge location, and de-identified timestamps of patient vitals data (pulse, blood pressure, temperature, blood oxygen saturation, and respirations), laboratory results, medications administered, amount, type, and route of fluid intake and output, neurologic assessments, and admission location. Data extraction and de-identification was performed by the institutional honest broker; study personnel had access only to this de-identified extracted data.

### Participant selection

Patients were included in the analysis sample if they were admitted to the hospital with a primary diagnosis of stroke, had a length of stay of at least 24 hours, and had at least one measurement of each: heart rate, blood pressure, and blood oxygen saturation documented in the first 24 hours of admission. Patients were excluded if they were admitted directly to a hospital unit as these patients were likely not admitted for acute stroke. Each patient encounter was treated independently; the same patient having multiple admissions for acute stroke captured in our dataset would be treated as two separate and otherwise independent patients.

### Processing

All data processing was performed using Python 3.8, notably relying on modules Pandas version 1.1.2 and NumPy version 1.18.5. Admission service was encoded as an admission to a neurology service versus a non-neurology service. Admission location was summarized as ICU (including neurological ICU), stroke unit (not including neurological ICU), or other. Admission time of day was summarized as a continuous variable between 0 and 1 with 0 representing 7:00 am, 0.5 representing 7:00 pm, and 1 representing 6:59 am. Ages greater than 90 were set equal to 90 to maintain de-identification, and race was categorized according to standard non-mutually exclusive categories. Length of hospital stay was calculated to the minute as the difference between the admission and discharge timestamps. This was then transformed into a binary variable representing a length of stay greater than or equal to 7 days or less than 7 days.

Comorbidities were categorized according to ICD-10 codes entered into the medical record at the time of admission then grouped into categories including hypertension, atrial fibrillation, cancer, diabetes, heart failure, kidney disease, and others. Ischemic and hemorrhagic strokes were similarly derived from manual grouping of ICD-10 codes. Medications were categorized according to type.

Vitals signs including pulse, blood pressure, oxygen saturation, temperature, and respiratory rate were extracted and clinically improbable values were removed, as these were believed to be errors in data entry. Time-dependent states such as patient location and the presence of IV catheters, drains, and airways were represented as time series functions representing cumulative time in each such state. For example, we constructed a time series variable for each patient capturing IV catheter presence where the value at each time point represents the total number of minutes spent with an IV catheter in pace up to that time. These time series variables are thus linearly increasing when the state is present and constant when the state is not present.

### Times series feature generation

Time series feature extraction was performed using Python module Tsfresh version 0.18.0. Summary statistics including mean, median, and entropy were generated from the time series data, along with higher order metrics such as Lempel-Ziv Complexity and Benford Correlation. Certain metrics could not be calculated due to sparsity of time series data; these metrics were removed. The combination of all features extracted is thought to be a comprehensive static representation of the time series data with minimal loss of information [6].

### Model building and statistical analysis

Given a national mean length of stay of 6.7 days for patients admitted to an ICU for stroke, we defined a prolonged length of stay as ≥ 7 days [7]. Descriptive statistics were evaluated to summarize the data sample stratified by LOS ≥ 7 days and < 7 days. Differences were assessed using t-tests for continuous variables and Chi-square tests for categorical and binary variables. To ensure against overfitting, the study sample was randomly split at the patient level into a training set (70%) and a testing set (30%). Models were trained and optimized only on data from the training set, and the testing set was used only for final performance assessment.

Using a binary outcome of prolonged length of stay vs not prolonged length of stay, logistic regression (GLM), random forest (RF), and XGBoost (XGB) models were created with a dataset composed of Tsfresh-generated features and other static data as predictors. Logistic regression models are highly interpretable and contain easily accessed parameters that can be useful for feature ranking. Similarly, random forest and XGB algorithms have established high levels of performance on comparable tabular data problems and are robust against overfitting.

The first model built utilized only static and hemodynamic data available during the first 24 hours of patient admission. This initial 24 hour block is considered the patient data observation window. The final feature space of this model, including static features and time-series derived features, was generated from this 24 hour window of raw data. Secondary models were also created in which the observation window of patient data was dynamically increased. Rather than use only the first 24 hours of available patient data, these models utilize the first 48 hours or 72 hours while still predicting binary prolonged length of stay from patient admission. For models using 48 and 72 hours of data, patients with LOS shorter than 48 or 72 hours were removed, respectively, resulting in fewer patients used in each subsequent model. This slightly altered the class imbalance of each additional prediction task.

The models were then independently optimized using random hyperparameter search with 5-fold cross validation. For logistic regression models, the sole hyperparameter tuned was “C” on a scale log [0.001: 100]. The “max_iter” parameter was set to 20, and all other parameters were set equal to their default sklearn values. For random forest models the following hyperparameters were optimized: “min_samples_split” was tuned over [1, 2, 3, 4], “min_samples_leaf” was tuned over [0.1, 0.3, 0.4, 1], “max_features” was tuned over [‘auto’, ‘sqrt’, ‘log2’]. All other parameters were set to sklearn defaults. For XGB models, the following hyperparameters were tuned: “booster” was tuned over [‘gbtree’, ‘gblinear’, ‘dart’], “eta” was tuned over [0.1, 0.2, 0.3, 1], “max_depth” was tuned over [4, 6, 8]. All other parameters were set to XGB defaults.

Subsequently, we employed feature ranking to optimize performance while reducing the size of the feature space, then repeated hyperparameter optimization. For the logistic regression model, all features were ranked based on the resulting coefficients using Lasso (L1) regression. Lasso regression favors a sparse feature space, so a majority of features received a coefficient of 0. To determine the updated feature space, all features with non-zero coefficients were retained. This new feature space was then used to retrain the logistic regression models. Unlike logistic regression, random forest and XGB are tree-based algorithms, so the Lasso-derived feature space would most likely not translate as well to these methods. To derive a unique feature space for random forest and XGBmodels, all features were ranked using random forest’s built-in feature importance metric. This calculates the importance of each feature by computing the average decrease in impurity across each split for each feature. This allows for the magnitudes of the resulting feature importance to be compared across the original feature space. By setting a threshold on this RF score, only features with scores above the selected threshold will be retained in the model. To optimize this threshold, a variety of thresholds were observed with their resulting feature spaces used to train a random forest classifier. This random forest feature space was also used for XGB models as they both use tree structures.

Performance of each model was assessed by calculating the area under the receiver operating characteristic curve (AUC). The mathematically optimal operating point was calculated as the point on the curve minimizing the Euclidean distance to the point (0,1) - that is, the point on the curve closest to the top left corner. This operating point was used to calculate sensitivity, specificity, accuracy, F1 score, positive predictive value, and negative predictive value for each model. Precision-recall (PR) curves were also plotted to observe the trade-off between positive predictive value and true-positive rate. These curves ignore true negative outcomes, so they are particularly informative for imbalanced datasets. All metrics were calculated using sklearn version 0.24.2.

## RESULTS

### Sample characteristics

Of 3,333 unique encounters, 2,025 met this study inclusion/exclusion criteria, representing 1,901 unique patients. The breakdown of patient enrollment can be seen in Figure 1. Table 1 shows sample characteristics by LOS. Approximately 35% (709/2,025) had a LOS ≥ 7 days. A higher percentage of patients in the LOS ≥ 7 days group were Black or African American compared to the LOS < 7 days group. Patients with LOS ≥ 7 days were also more likely to have hypertension, diabetes, atrial fibrillation, heart failure, and kidney disease. Approximately 70% (1413/2025) of patients in our sample had ischemic strokes vs hemorrhagic strokes. Patients with LOS ≥ 7 days were more likely to have hemorrhagic strokes than ischemic compared to patients with LOS < 7 days.

**Figure 1.**
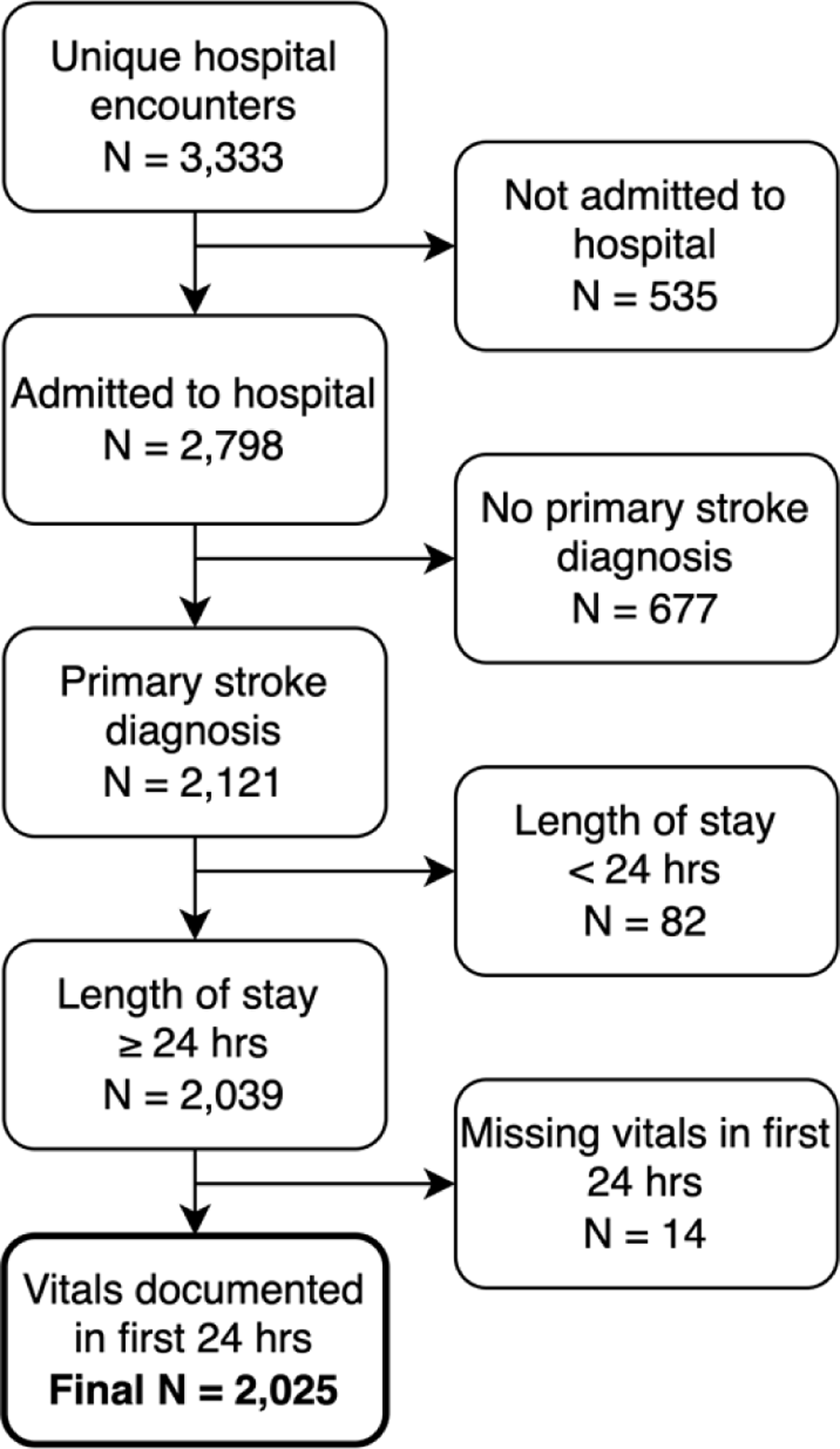
Inclusion/Exclusion criteria.

**Table 1.** Characteristics of sample based on prolonged length of stay.

| | Overall<br>n= 2025 | LOS ≥ 7 days<br>n= 709 | LOS < 7 days<br>n=1316 | P-Value |
| --- | --- | --- | --- | --- |
| Age, mean (SD) | 64.4 (15.3) | 63.4 (15.1) | 64.9 (15.4) | 0.034 |
| Woman, n (%) | 1007 (49.7) | 354 (49.9) | 653 (49.6) | 0.931 |
| Race |  |  |  |  |
| White, n (%) | 1086 (53.6) | 334 (47.1) | 752 (57.1) | <0.001 |
| Black or African American, n (%) | 787 (38.9) | 306 (43.2) | 481 (36.6) | 0.004 |
| Asian, n (%) | 51 (2.5) | 25 (3.5) | 26 (2.0) | 0.048 |
| Other, n (%) | 108 (5.3) | 52 (7.3) | 56 (4.3) | 0.005 |
| Medical History |  |  |  |  |
| Hypertension, n (%) | 600 (29.6) | 276 (38.9) | 324 (24.6) | <0.001 |
| Diabetes, n (%) | 241 (11.9) | 106 (15.0) | 135 (10.3) | 0.002 |
| Atrial fibrillation, n (%) | 94 (4.6) | 50 (7.1) | 44 (3.3) | <0.001 |
| Heart failure, n (%) | 67 (3.3) | 35 (4.9) | 32 (2.4) | 0.004 |
| Kidney disease, n (%) | 75 (3.7) | 45 (6.3) | 30 (2.3) | <0.001 |
| Cancer, n (%) | 138 (6.8) | 56 (7.9) | 82 (6.2) | 0.184 |
| Stroke type, n (%) |  |  |  | <0.001 |
| Hemorrhagic | 612 (30.2) | 333 (47.0) | 279 (21.2) |  |
| Ischemic | 1413 (69.8) | 376 (53.0) | 1037 (78.8) |  |
| Admission time of day (time of day, | 17:54 (6.4) | 17:50 (6.4) | 18:00 (6.4) | 0.547 |
| hours), mean (SD) |  |  |  |  |

### 24-hour model

Tsfresh extraction on time series data up to 24 hours after admission yielded 8,741 non-zero features. Feature ranking, iterative removal, and repeat hyperparameter optimization were performed. ROC and precision-recall curves for this model are included in Figure 2. For GLM, feature ranking involved removing sparse features. For RF and XGB, feature ranking involved listing RF-derived feature importances and optimizing for the best threshold. This resulting feature space represented approximately 10% of the original features; this proportion appeared to consistently provide strong results when optimizing the number of features to retain when using feature ranking. Top scoring features, categorized by variable of origin, are shown in Figure 3. These results yielded test set ROC AUCs of 0.83, 0.82, and 0.85 for the GLM, random forest, and XGB models respectively. Additionally, these models scored corresponding precision-recall AUCs of 0.75, 0.74, and 0.78. While all three models performed well, the XGB model produced the highest AUC. The optimal operating point metrics for all models are listed in Table 2. The XGB model attained an accuracy of 80%, specificity of 81%, and sensitivity of 78%.

**Figure 2.**
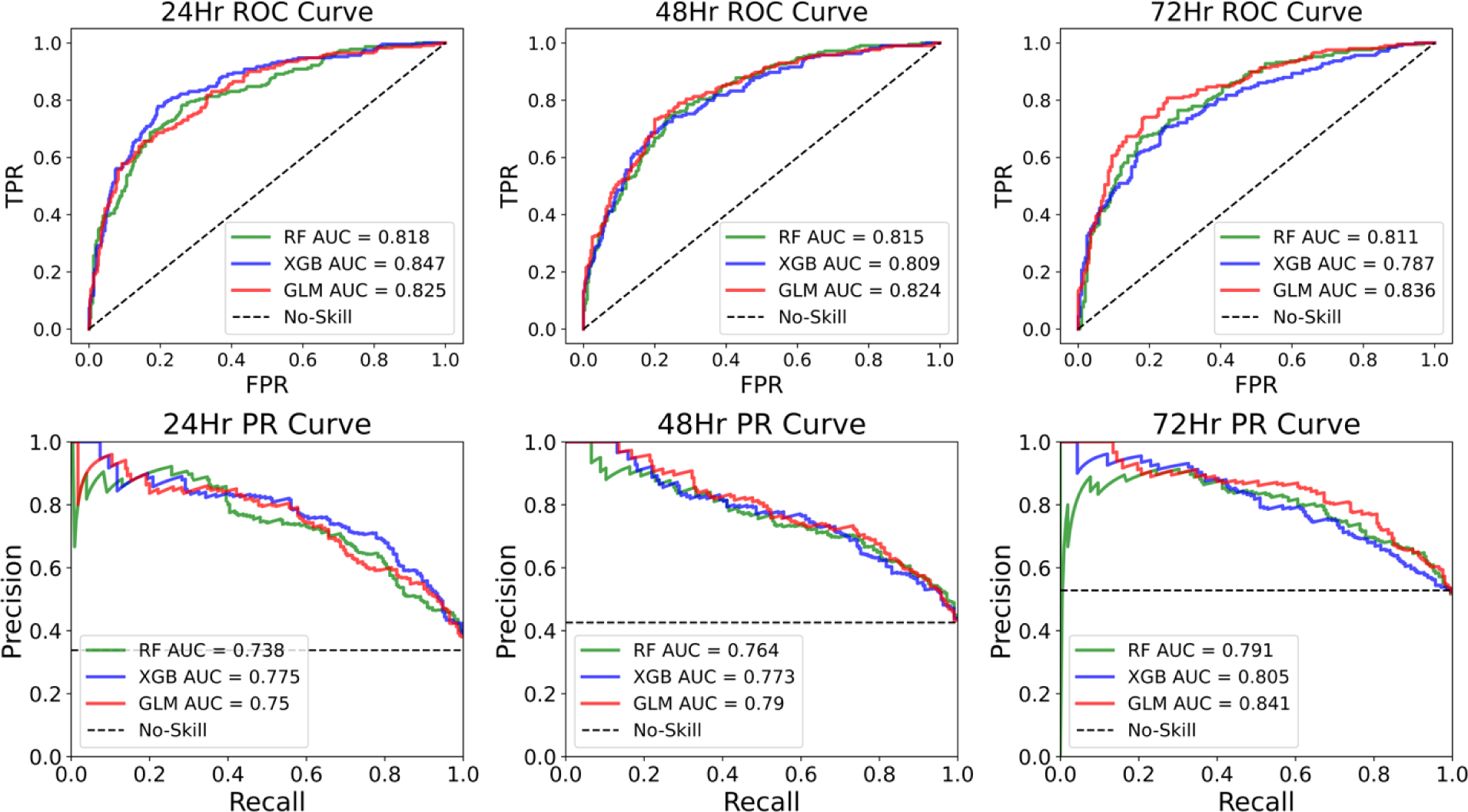
ROC and precision-recall curves for 24 hour, 48hr, and 72hr models. For the ROC curves (top), the y-axis represents true positive rate (TPR) and the x-axis represents false positive rate (FPR). For the precision-recall curves (bottom), the y-axis represents precision and the x-axis represents recall. All three algorithms are plotted on each figure, differentiated by color. The dotted black line represents the expected performance of a no-skill classifier. From these results, it appears XGBoost performs the best in 24hr models, and GLM performs best in 48 and 72hr models.

**Figure 3.**
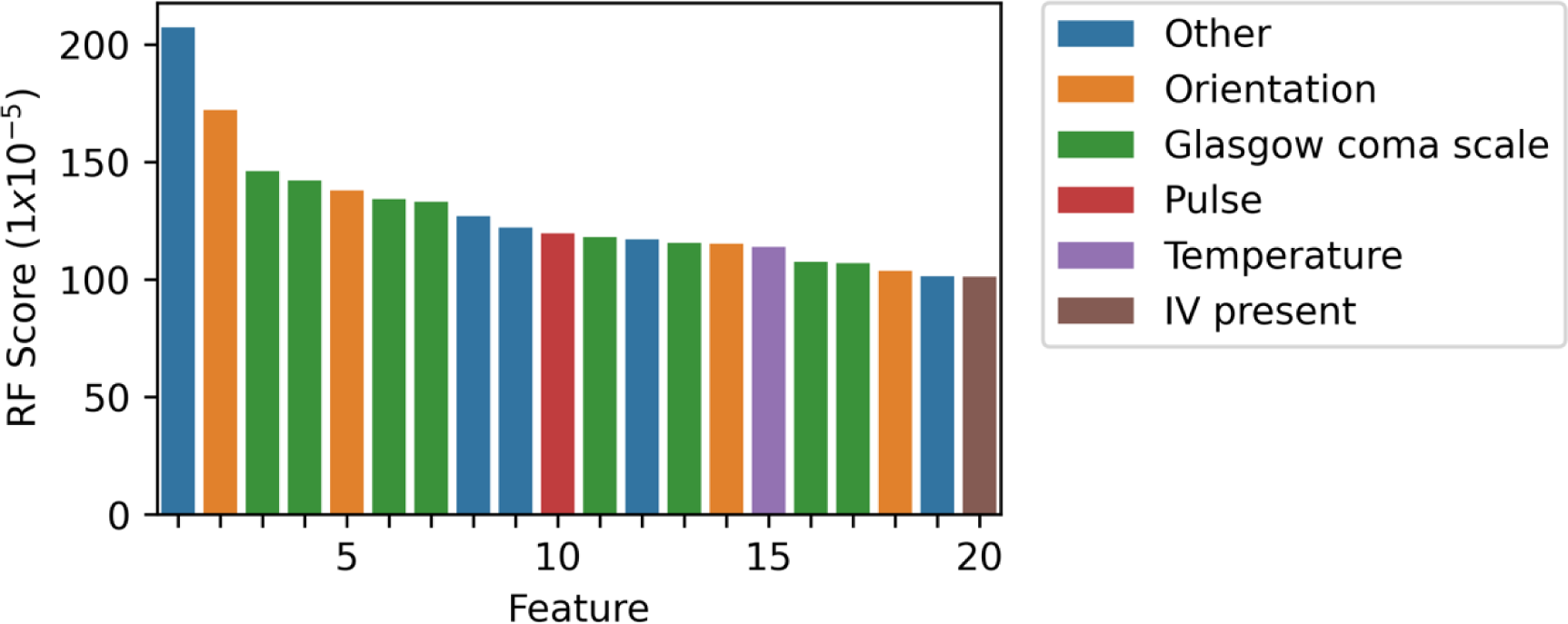
Top scoring features using random forest feature importance. Y-axis represents the attributed RF score of a given feature; x-axis represents the feature itself. Features derived from different categories of raw data are represented by different colors. GCS-derived features appear to have the most prevalence amongst top scores.

**Table 2.** Evaluation metrics for all models at each respective optimal operating point. For the 24 hour models, XGBoost performed best; for the 48 hour models, GLM performed best; for the 72 hour models, GLM performed best.

|  | AUC | Specificity | Sensitivity | PPV | NPV | F1-Score | Accuracy |
| --- | --- | --- | --- | --- | --- | --- | --- |
| 24hr (GLM) | 0.825 | 0.81 | 0.69 | 0.68 | 0.81 | 0.69 | 0.76 |
| 24hr (RF) | 0.818 | 0.74 | 0.78 | 0.65 | 0.85 | 0.71 | 0.75 |
| 24hr (XGBoost) | 0.847 | 0.81 | 0.78 | 0.71 | 0.86 | 0.74 | 0.80 |
| 48hr (GLM) | 0.824 | 0.80 | 0.73 | 0.73 | 0.80 | 0.73 | 0.77 |
| 48hr (RF) | 0.815 | 0.76 | 0.75 | 0.70 | 0.80 | 0.72 | 0.75 |
| 48hr (XGBoost) | 0.809 | 0.75 | 0.74 | 0.69 | 0.79 | 0.71 | 0.75 |
| 72hr (GLM) | 0.836 | 0.75 | 0.81 | 0.77 | 0.79 | 0.79 | 0.78 |
| 72hr (RF) | 0.811 | 0.72 | 0.76 | 0.74 | 0.75 | 0.75 | 0.74 |
| 72hr (XGBoost) | 0.787 | 0.76 | 0.71 | 0.75 | 0.71 | 0.73 | 0.73 |

### 48 and 72-hour models

The 48 and 72-hour models utilized data from 1,162 and 949 patient encounters respectively. Similar to the 24-hour model, initial models on the entire Tsfresh-generated feature space were optimized, then feature ranking and reduction followed by repeat hyperparameter optimization were used to arrive at final 48 and 72-hour models. ROC and PR curves for these models are also included in Figure 2. Operating point metrics are included in Table 2. GLM had the best AUC at both time points, at 0.82 and 0.84 for the 48 and 72-hour models respectively.

**GCS models**

## DISCUSSION

Our models predicted a cohort of patients with prolonged length of stay using the first 24 hours of clinical data in a large population of hospitalized stroke patients. Our best model to predict prolonged length of stay (LOS ≥ 7 days) using data from the first 24 hours of admission was an XGB model that achieved an AUC of 0.85 with accuracy of 80% at the operating point, demonstrating a strong ability to predict who will have a long LOS after only the first day, which could enable selection of patients for early coordination of care and planning for patients and their care partners. It additionally suggests that we can identify a cohort of stroke patients who will have a longer recovery trajectory. Clinicians can potentially use this risk profile to implement changes to a patient’s therapy plan which could improve outcomes.

By examining the top ranked features for the 24 hour XGB model, we found that many GCS time-series features populated the top 20 most important features. Although we expected hemodynamic features to rank highly, they were scarce in the top 20 features. When we expanded the range to the top 100 ranked features, we saw that hemodynamic parameters were well represented, but no specific hemodynamic features (e.g., pulse, blood pressure) dominated the most important features. This suggests that while no specific hemodynamic features were of singular importance, they likely played an important role en masse in predicting the binary length of stay outcome.

By repeating our modeling process using data up to 48 and 72-hours of admission, we were able to evaluate model performance based on varying amounts of data. While we initially expected model performance to increase with more data, we largely observed unclear patterns over 24, 48, and 72-hour models. The exception is the XGB architecture, where we observed a decrease in AUC, sensitivity, and specificity with increasing amounts of data. These patterns suggest that there may be confounding issues comparing across time frames, since each model (24, 48, 72 hours) had a different subsample of the training data. For example, the training data for the 48-hour model screens out patients admitted for less than 48 hours whereas these patients were included in the 24-hour model. This results in shifting class imbalances between these models, gradually favoring the prolonged LOS outcome as the time window increases. Thus, the prevalence of different patient characteristics--including our length of stay outcome--changes between models. This changing subsample may be confounding the ability to directly compare these models across time. Future work is needed to test whether this is indeed the case or if there is another factor limiting the performance of models with increasing data.

Past research has sought to predict length of hospital stay and outcomes of stroke patients. Using only parameters available at admission for 330 ischemic stroke patients, Chang et al. performed multiple regression analysis with five parameters selected by univariate analysis [8]. Using sex, smoking status, NIH Stroke Scale score, Modified Barthel Index score, and a binary indicator for whether the stroke was a small-vessel occlusion, the model achieved an R^2^ of 0.369. Naito et al. investigated the relationship between blood pressure during the subacute phase of ischemic stroke and 3-month functional outcome [9]. While the average blood pressure had no association with outcome, the standard deviation of systolic and diastolic pressure and coefficient of variation of diastolic pressure were independently associated with poor outcome at 3 months. This study did not report the LOS of the patients. Compared to past research, these models have particularly strong performances [8]. Since all three LR, RF, and XGB models performed roughly equivalently, the strength of our results would appear to be in the feature space. While past research has focused largely on static data available at the time of admission or shortly after, we were able to include an array of time series features including vitals, neurological assessments, lab results, and medication administrations [7]. Whereas past research has largely used continuous outcomes, stratifying into a meaningful binary threshold may have also contributed to the high performance of our models [8].

This work has several important limitations. The sample population was admitted to a single site, and the specific care pathways of this single institution likely impact length of stay, limiting the generalizability of our findings. That said, because acute stroke care is driven by guidelines in the first week, the results may be generalizable to other urban comprehensive stroke centers. Further exploration of our models at different clinical sites may assist in supporting generalizability. Another limitation is the existence non-medical factors that may artificially impact hospital length of stay, such as delays at the discharge location, insurance plan nuances, and social situations for which we were unable to measure and control.

In all, this work presents preliminary tools to predict prolonged length of stay for hospitalized stroke patients. Our hope is that with further refinement these tools may be used in the clinical space to indicate potential for complications in the early stroke recovery course, assist in activating early and efficient discharge planning to maintain continuity of care, as well as serve as an early indicator that may trigger additional interventions if a patient is predicted to have a poor outcome.

## SUMMARY

By using existing clinical data during routine stroke admission, we were able to predict, based on only the first 24 hours following admission, whether patients will have a prolonged length of stay with an AUC of 0.85. Future work is needed to refine and optimize these models for implementation in the clinical space, where they may be used to facilitate efficient discharge planning as well as an indicator of patients with risk profiles suggestive of poor outcomes.

## Data Availability

The datasets generated during and/or analyzed during the current study are available from the corresponding author on reasonable request.

## Acknowledgement

The authors would like to acknowledge the work of Alex Hepp, Varun Naga, MSE, Athena Olszewski on this project and funding from the American Heart Association Innovative Project Award.

## NON-STANDARD ABBREVIATIONS AND ACRONYMS

AUC: Area under receiver operating characteristic curve
EHR: Electronic health record
FPR: False positive rate
GCS: Glasgow Coma Scale
GLM: Generalized linear model
LOS: Length of stay
LR: Logistic regression
PR: Precision-recall
RF: Random forest
ROC: Receiver operating characteristi
TPR: True positive rate
XGB: XGBoost

